# Proteome-wide and mendelian randomisation analyses of chronic widespread pain: diagnosis, prognosis, and drug target discovery

**DOI:** 10.1101/2024.10.29.24316353

**Authors:** Li Chen, Eoin Kelleher, Ruogu Meng, Duanke Liu, Yuchen Guo, Yunhe Wang, Yaoqing Gao, Zhe Huang, Zhu Liang, Shuai Yuan, Chao Zeng, Guanghua Lei, Jun Ma, Yanhui Dong, Anushka Irani, Junqing Xie, Daniel Prieto-Alhambra

## Abstract

Chronic widespread pain (CWP) remains challenging due to its heterogenous causes and complex mechanisms. We analysed 2,923 plasma proteins from 29,254 UK Biobank participants. We first identified 811 proteins correlated with the presence of CWP cross- sectionally. We then created a sparse (top 10 proteins) and intricate (all significant proteins) proteomic-based score (ProtS) for CWP, both outperforming and improving the traditional clinical score (AUC: 0.82 and 0.88 vs 0.81 individually, and 0.87 and 0.92 in combination). Prospectively, the ProtS was associated with increased risks of a spectrum of pain traits, including dimensions from pain onset, progression and intensity, up to 13-years of follow-up; More importantly, we identified distinct proteomic signatures for nociplastic pain compared to nociceptive and neuropathic pain. For the individual proteins, carbonic anhydrase 14 (CA14) and leptin appeared as promising casual CWP biomarkers as triangulated by Mendelian randomisation and colocalization analyses. Lastly, our drug-repurposing analysis identified ten potential candidates for CWP therapies, calling for further research including randomised controlled trials. Although no CA14 agonists are currently available, CA14 remains a promising target, warranting further efforts to explore its role in pain modulation.

## Introduction

Chronic pain affects 27.5% of the global population and is one of the leading causes of medical consultation and disability^1,2^, particularly among adults over 65, where joint, back, and neck pain are common ^3,4^. The burden of chronic pain extends beyond physical suffering, as it poses signifanct societal costs through lost productivity and increased healthcare resource utilisation, presenting a major global health challenge^5,6^.

The International Association for the Study of Pain (IASP) defines pain as an unpleasant sensory and emotional experience. While acute pain serves a protective function by signaling potential or actual tissue damage, chronic pain is defined as pain persisting for more than three months, continuing beyond the normal healing period. Unlike acute pain, which is adaptive, chronic pain is often maladaptive as it no longer serves a protective role. Indeed, chronic pain is increasingly recognised as a distinct condition, with the 2019 ICD-11 classification introducing ‘chronic primary pain’, where pain is the principal problem rather than a result of another disease ^7–10^. Chronic pain was traditionally classified as either neuropathic pain, which originates from a lesion or disease of the somatosensory nervous system, or nociceptive pain, which results from actual or threatened non-neural tissue damage^8,9^. However, in 2016, a third category—nociplastic pain—was introduced to describe pain arising from altered nociception without clear evidence of tissue or nerve damage. Nociplastic pain, characterised by its widespread and diffuse nature, is particularly debilitating and is best typified by conditions such as fibromyalgia^11^.

Despite advances in understanding these pain mechanisms, clinical tools to accurately diagnose and distinguish between them remain lacking. Existing clinical models do not account for the molecular underpinnings that could differentiate nociceptive, neuropathic, and nociplastic pain. Here, proteomics – the large-scale study of proteins – offers a promising avenue. By examining proteins as biomarkers, proteomics offers promising opportunities to improve diagnostic accuracy and support personalised therapeutic strategies for chronic pain.

The field of proteomics has significantly advanced our understanding of molecular mechanisms in complex diseases like cardiovascular disease, diabetes, and cancer by integrating genetic, lifestyle, and environmental influences ^12^. Given the recognition of pain vulnerability—where genetic and biological factors predispose certain individuals to developing chronic pain^13–15^—proteomic analyses could help identify those at higher risk, facilitating early recognition and intervention. Prior biomarker studies in chronic pain have been limited by small sample sizes and a narrow focus on specific proteins, failing to capture the full complexity of pain mechanisms ^16^. By leveraging a comprehensive proteomic dataset in a large population-based study, our research addresses this gap and provides an opportunity to identify novel biomarkers associated with pain vulnerability and distinct pain mechanisms^17^. These innovative approaches may offer a deeper understanding of chronic pain mechanisms, opening pathways to improved diagnostic assays and novel therapeutic targets.

We therefore aimed to: (1) evaluate the proteomic signatures distinguishing acute and chronic pain, identifying unique molecular patterns; (2) assess the diagnostic and prognostic validity of models based on 2,923 unique plasma proteins in predicting widespread chronic pain, comparing their performance to conventional clinical risk models, and to explore the added value of integrating plasma proteins into clinical risk models; and (3) to assess the causal relevance of key identified proteins and their potential as therapeutic targets for the treatment of chronic pain.

## Results

### Baseline Characteristics and Pain Phenotypes

We analysed 29,254 participants with plasma proteomics data (Olink) from the UK Biobank, of whom 44.6% were male, with a median age of 55.8 years (**Supplementary Table 2**). The comparison of participants with and without proteins is provided in **Supplementary Table 1**. In the present study, 52.3% of participants reported suffering chronic pain at baseline. Regarding pain types, 85.2% reported musculoskeletal pain, 22.5% pain in the head and face, 10.7% abdominal pain, and 3.0% experienced widespread pain. Overall, 24.4% of participants reported at least two pain sites affected. A Venn diagram depicting the overlap between different chronic pain types, including widespread chronic pain, is provided in **Extended Data Fig. 1**. In 2019, data on pain mechanisms were collected from a total of 8,225 participants. Among them, 571 reported nociceptive pain, 75 reported neuropathic pain, 4,153 experienced nociplastic pain, and 717 had a diagnosis of fibromyalgia (**Supplementary Table 2)**.

### Proteomic Associations with Chronic and Acute Pain

We conducted a proteome-wide association study (PWAS) to evaluate the ability of protein signatures to distinguish chronic pain by comparing protein expression profiles between individuals with chronic pain and pain-free controls. People with chronic pain showed widespread and substantial changes in protein expression compared to pain-free controls. In contrast, those with acute pain showed minimal associations with protein levels **(Fig.1A and Extended Data Fig. 2**). To assess the difference in protein signatures associated with chronic and acute pain, we analysed the overlap between proteins linked to each condition. Only 27% (234/864) of the proteins associated with chronic pain overlapped with those linked to acute pain, underscoring distinct molecular pathways (**Fig.1A and B and Supplementary Table 3**).

**Figure 1.**
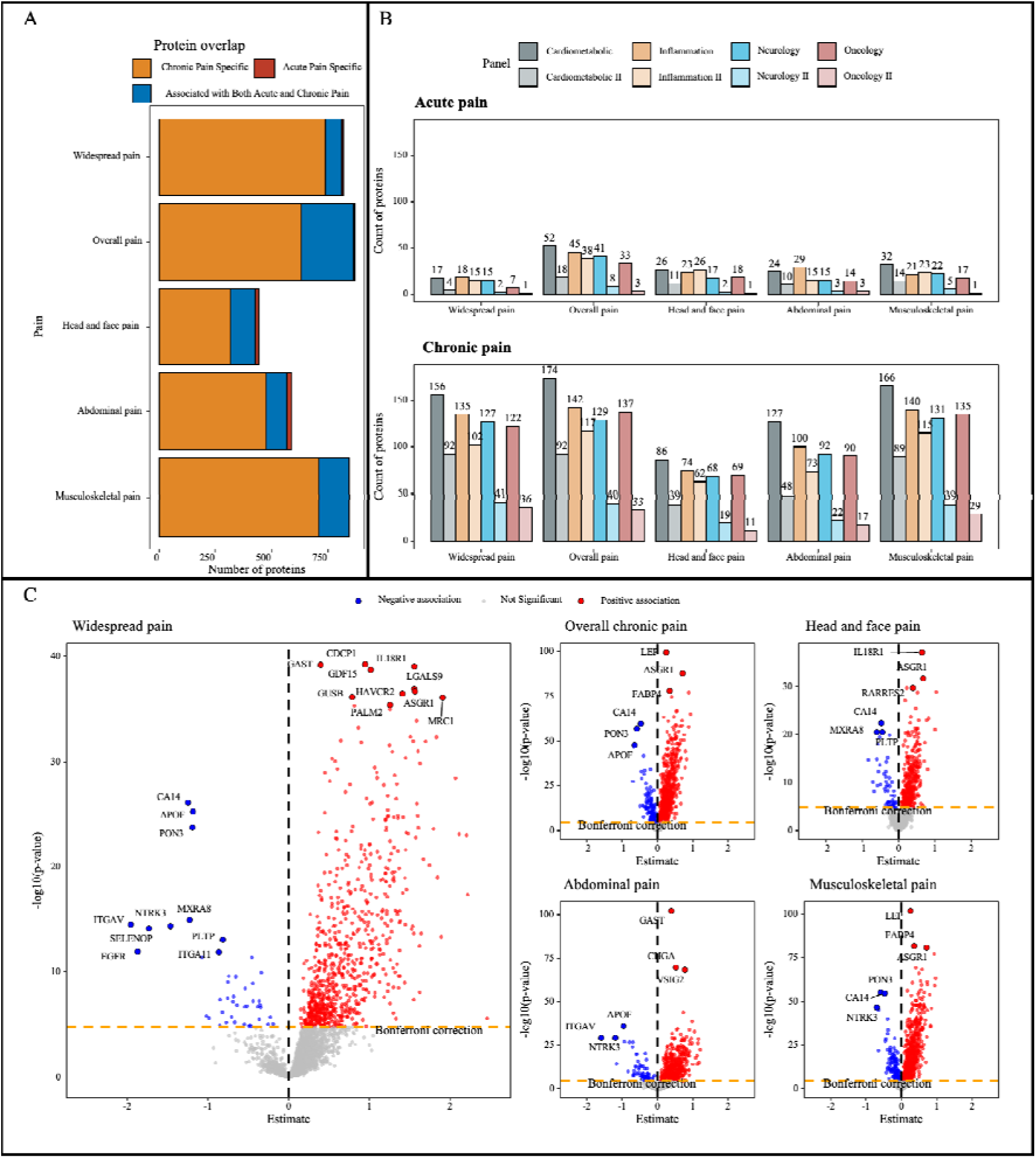
Proteomic signature of different types of chronic pain and acute pain. Notes: A, Comparison of proteins significantly associated with both chronic and acute pain. B, Count of proteins significantly associated with specific types of acute and chronic pain, categorised by panel. C, Volcano plots show protein associations with different pain types after applying Bonferroni correction. Proteins with p-values below 0.05/2923 are highlighted in red (positive) and blue (negative), while nonsignificant associations are represented in gray. The top 10 positively and negatively associated proteins with the lowest p-values for each pain type are labelled.

In the analysis of widespread chronic pain, the proteins most significantly upregulated compared to controls were CDCP1, IL18R1, GAST, GDF15, LGALS9, GUSB, ASGR1, MRC1, PALM2, and HAVCR2. In contrast, the ten most downregulated proteins were CA14, APOF, PON3, NTRK3, MXRA8, ITGAV, SELENOP, PLTP, EGFR, and ITGA11 (**Fig.1B**). A total of 811 proteins out of 2,923 (27.7%) exhibited significant associations with widespread chronic pain. In the PWAS, 864 out of 2,923 proteins (29.6%) exhibited significant associations with overall chronic pain. Musculoskeletal pain had the highest number of significant associations, with 844 proteins (28.9%), followed by abdominal pain with 569 proteins (19.5%), and head and face pain with 428 proteins (14.6%) (**Fig. 1B and Supplementary Table 4**). Several proteins, including APOF, ASGR1, CA14, CA6, MXRA8, PLTP, PON3, and RARRES2, were shared across multiple pain types (**Fig. 1C**).

### Intricate and sparse proteomic scores (I-ProtS and S-ProtS)

Proteins identified from PWAS for widespread chronic pain and specific chronic pain types were used to derive intricate proteomic scores (I-ProtS) for chronic pain. Additionally, sparse proteomic scores (S-ProtS) were generated using the top 10 proteins associated with widespread chronic pain and specific chronic pain types to evaluate the potential of these sparse plasma protein signatures in distinguishing specific chronic pain types.

Higher baseline percentiles of both I-ProtS and S- ProtS correlated with increased prevalence of chronic pain at baseline, both across overall as well as for specific chronic pain types. The associations between I-ProtS and overall chronic pain are illustrated in **Extended Data Fig. 3**, showing a strong correlation coefficient [95% CI]: 0.99 [0.99–1.00]. Across all chronic pain types, S-ProtS showed a distribution similar to I-ProtS and comparable correlations with observed event rates. The association between I-ProtS or S-ProtS and the observed rates of widespread pain differed from that of other specific pain types (**Extended Data Fig. 3**).

### Protein signatures improved classification of widespread chronic pain

Clinical risk models (CS) incorporating previously^13^ identified key risk factors like sleep difficulties, neuroticism, powerlessness, mental health issues, life stressors, and obesity achieved a median Area Under the Curve (AUC) of 0.81 (95% CI: 0.76–0.86) for widespread chronic pain. A protein-only model based on S-ProtS showed comparable performance, with an AUC of 0.82 (95% CI: 0.77–0.87). The I-ProtS model achieved numerically higher performance, with an AUC of 0.88 (95% CI: 0.83–0.92). Incorporating I-ProtS into the CS model enhanced discrimination, yielding an AUC of 0.87 (95% CI: 0.83–0.91) for widespread chronic pain. The combined I-ProtS and CS model reached the highest discrimination, with an AUC of 0.92 (95% CI: 0.88–0.94) (**Fig. 2A**). To assess the interpretability of the classification models, we evaluated their calibration and performed decision curve analyses. For widespread chronic pain, the combined models demonstrated good calibration, as evidenced by a calibration slope of 0.97 (95% CI 0.85–1.10), indicating strong alignment between predicted and observed risks (**Extended Data Fig. 4**.).

**Figure 2.**
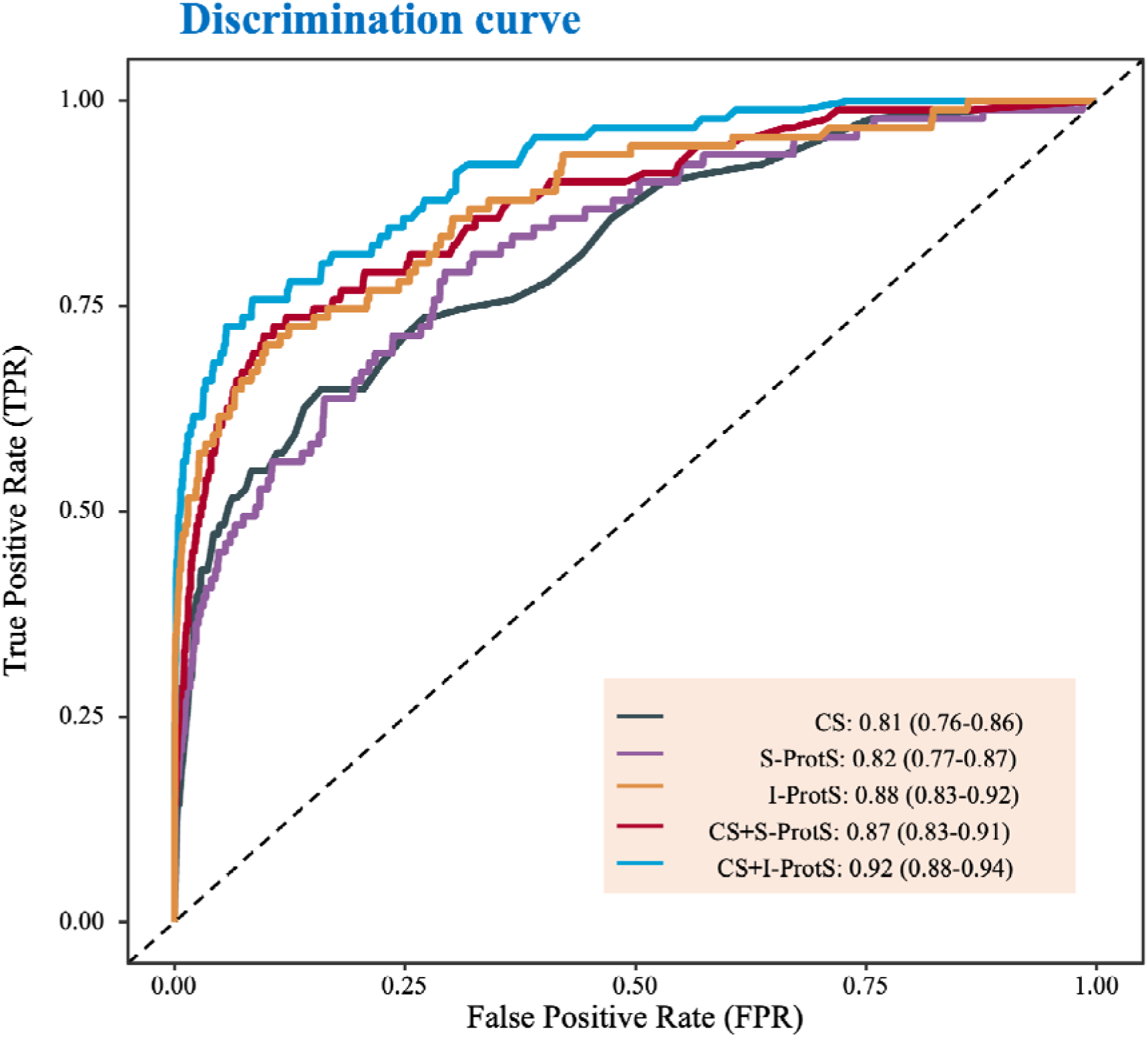
Diagnostic performance of proteomic score, clinical score, and combined. **Notes**: this figure presents the ROC curves and Area Under the Curve (AUC) for widespread chronic pain. Higher AUC values indicate improved model performance in distinguishing individuals with widespread chronic pain from those without. **Abbreviations**: CS: clinical risk model; S-ProtS: sparse proteomic score; I-ProtS: intricate proteomic score; CS+S-ProtS: combined model with clinical model and S-ProtS; CS+I-ProtS: combined model with clinical model and I-ProtS.

CS models for chronic pain had AUC ranging from 0.66 to 0.76, with the highest performance observed for abdominal pain (AUC = 0.76, 95% CI: 0.73–0.79). Across the three pain types, the I-ProtS demonstrated comparable performance to the CS model. However, the sparse proteomic score (S-ProtS) model did not achieve superior or comparable performance relative to the CS model for any chronic pain types (**Extended Data Fig. 4**). Incorporating S-ProtS into the CS model significantly enhanced discrimination for all chronic pain types, with an AUC of 0.69 (95% CI: 0.67–0.70) for Musculoskeletal pain and the best performance for abdominal pain (AUC = 0.81, 95% CI: 0.78–0.83). The addition of I-ProtS to the CS model further improved classification performance beyond that of S-ProtS, with an AUC of 0.91 (95% CI: 0.90–0.92) for head and face pain (**Extended Data Fig. 4**). The I- ProtS-only model showed a net benefit comparable to that of the CS-based models, while the combined models (incorporating I-ProtS or S-ProtS) provided a greater net benefit than models based solely on either CS or I-ProtS alone (**Extended Data Fig. 4**).

### Prospective Prediction of Pain Progression and Onset

The stability and individual changes in pain site numbers between baseline and the two follow-up visits were shown in **Extended Data Fig. 5**. We assessed the predictive power of I- ProtS and S-ProtS for predicting new widespread pain, pain mechanisms, and pain progression. Both I-ProtS and S-ProtS demonstrated good predictive discrimination for fibromyalgia, with AUCs of 0.76 and 0.74, respectively (**Fig. 4A**). The ProtS showed better calibration for the nociplastic pain and fibromyalgia (**Supplementary Fig.1**).

**Fig. 4:**
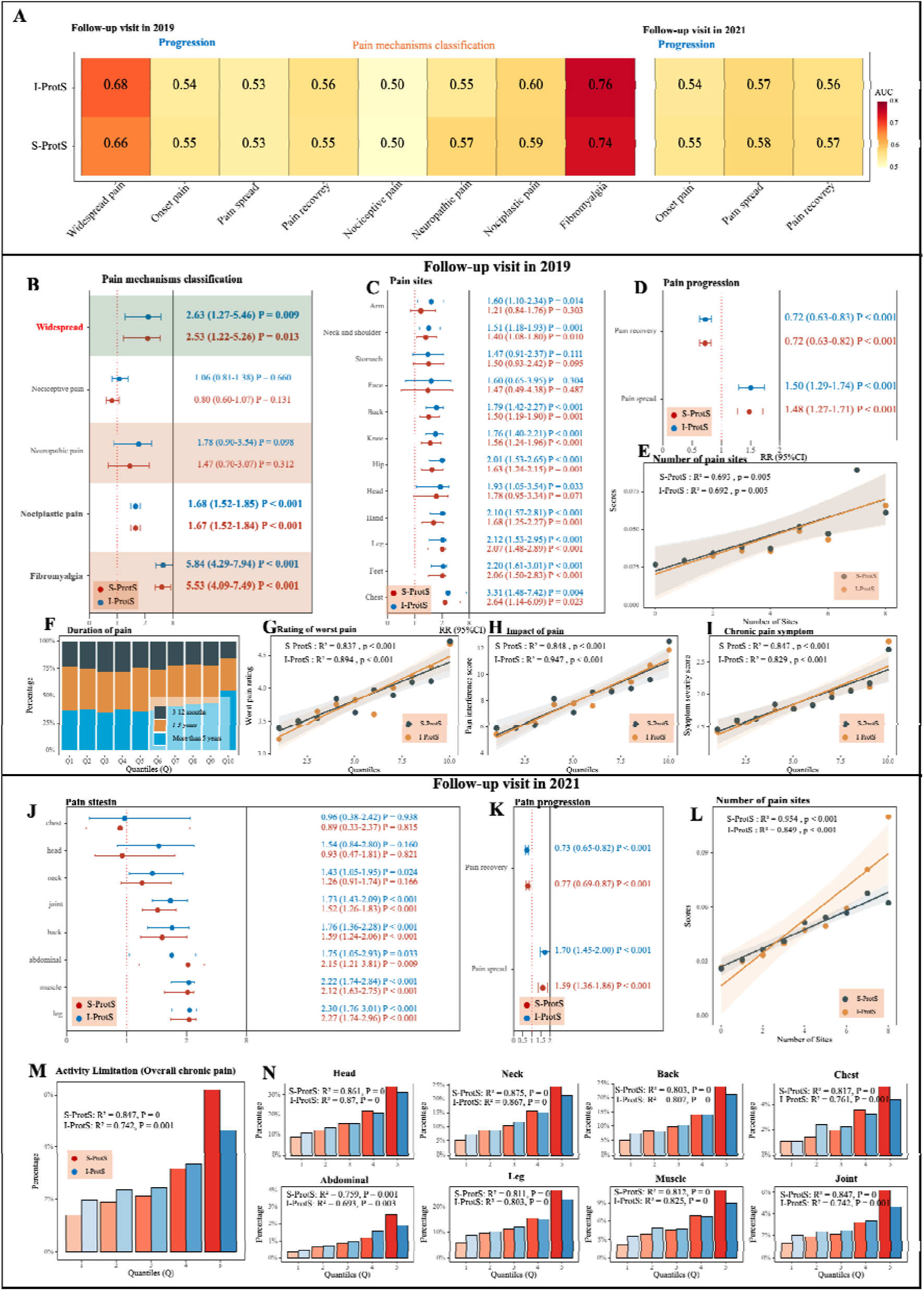
Prospective associations of pain-specific proteomic scores with mechanism pain classification, pain onset, progression, multisite chronic pain, duration, intensity, impact, and activity limitations in longitudinal data. Notes: The 2019 follow-up refers to the online follow-up conducted in 2019, and the 2022 follow-up refers to the one conducted in 2022. Panel A shows the AUC of I-ProtS and S- ProtS for selected pain types. Panel B: Displays the association between the onset of widespread chronic pain, pain mechanism classification, and the proteomic scores (ProtS) in 2019. Panels C and J: Show the associations between the onset of chronic pain and the proteomic scores in 2019 and 2022, respectively. “Onset of chronic pain” includes participants who were pain-free at baseline but reported chronic pain at follow-up. Panels D and K: “Pain spreading” refers to participants whose number of pain sites increased, while “pain recovery” refers to those whose pain sites decreased. Panels E and L: Depict the number of pain sites (multisite chronic pain) plotted against quantiles of the Sparse Proteomic Score (S-ProtS) and Intricate Proteomic Score (I-ProtS). Panel F: Shows the proportion of chronic pain duration across quantiles of I-ProtS. Panels J-I: Illustrate the rating of worst pain, impact of pain, and symptom severity, respectively, in relation to quantiles of S-ProtS and I- ProtS. Panel M: Illustrates the proportion of activity limitations caused by overall chronic pain among participants with chronic pain. Panel N: Shows activity limitations by specific chronic pain sites. Abbreviations: S-ProtS, Sparse Proteomic Score; I-ProtS, Intricate Proteomic Score.

Both I-ProtS and S-ProtS for widespread chronic pain were significantly associated with the onset of widespread chronic pain at follow-up visits (**Fig. 4B**). In the pain mechanism classification, I-ProtS and S-ProtS were significantly associated with nociplastic pain and fibromyalgia. However, for nociceptive pain and neuropathic pain, no significant associations were observed for either I-ProtS or S-ProtS (**Fig. 4B**).

For the onset of specific pain sites, I-ProtS and S-ProtS also showed significant positive associations, although these were weaker for certain pain sites, such as headache, facial pain, and abdominal pain, compared to other chronic pain sites (**Fig. 4C** **and J**). Both follow-up visits demonstrated a positive correlation between proteomic scores and pain spread, and a negative correlation with pain recovery. Higher proteomic scores were associated with an increased number of pain sites (multisite chronic pain), while lower scores corresponded with fewer pain sites and greater pain recovery (**Fig. 4D, E, K, and L**). The proteomic scores, both I-ProtS and S-ProtS, demonstrated a strong association with the number of pain sites at the 2019 follow-up (I-ProtS: R^2^= 0.692, P=0.005; S-ProtS: R^2^=0.693, P=0.005) and an even stronger association at the 2022 follow-up (I-ProtS: R^2^=0.849, P<0.001; S-ProtS: R^2^=0.954, P<0.001) (**Fig. 4C** and **G**).

Proteomic scores exhibited a monotonic increase with pain duration, rating of worst pain, pain impact, and pain symptom severity nine years later (**Fig. 4F-I**). The percentage of participants reporting activity limitations due to chronic pain increased across higher proteomic score quantiles (I-ProtS and S-ProtS) (**Fig. 5L**). S-ProtS showed comparable associations with activity limitations related to chronic pain sites, similar to those observed with I-ProtS (**Fig. 4N**).

**Fig. 5:**
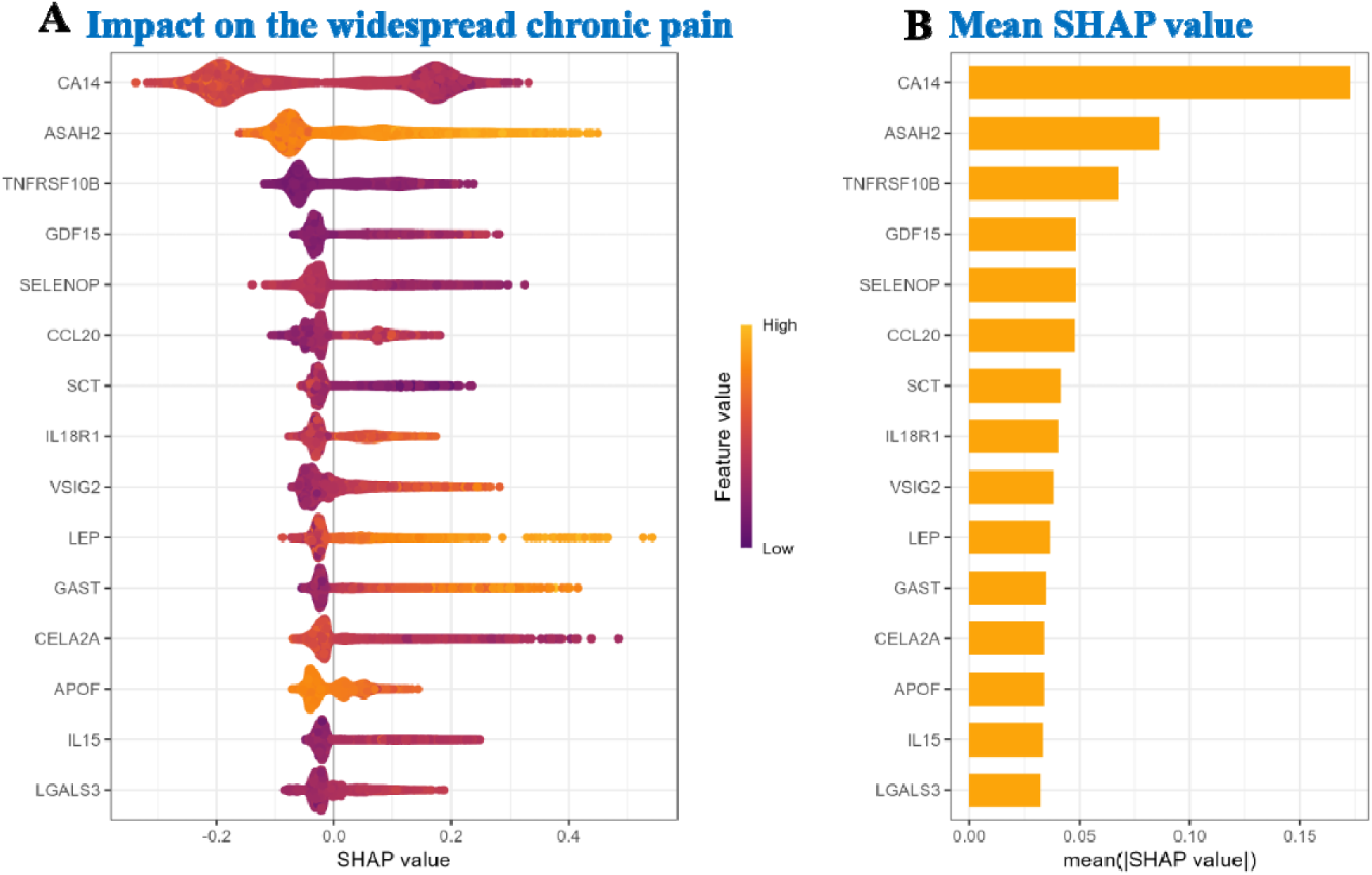
SHAP of protein contributions to widespread chronic pain. Notes: (A) SHAP plot illustrating the impact of individual proteins on widespread chronic pain prediction. Brighter colours (yellow) represent higher feature values, while darker colours (purple) indicate lower feature values, demonstrating how each protein contributes to the prediction model. (B) Mean SHAP values for the top contributing proteins to widespread chronic pain. Abbreviations: SHAP, SHapley Additive exPlanations.

### Contributions of proteins to classification of widespread chronic pain

Proteins including CA14, ASAH2, TNFRSF10B, GDF15, SELENOP, CCL20, SCT, IL18R1, VSIG2, LEP, GAST, CELA2A, APOF, IL15, and LGALS3 were identified as key contributors to widespread chronic pain. Among these, CA14 emerged as the most impactful, reflected by its highest SHAP (SHapley Additive exPlanations) value, suggesting a prominent role in pain prediction. ASAH2 and TNFRSF10B followed as major contributors. The SHAP analysis further highlighted the role of LEP, where both high and low levels influenced risk prediction. Elevated LEP values (indicated by yellow) are particularly associated with an increased risk of widespread chronic pain (**Fig. 5**).

SHAP analyses identified several key proteins contributing to the classification of different chronic pain types. LEP consistently emerged as the top predictor, exerting a strong influence on the model. Other proteins, including COL9A1, GAST, SELENOP, and CRTC1, also displayed high SHAP values, underscoring their roles in pain prediction. COL9A1 and GAST demonstrated particularly notable impacts. In specific pain types, proteins such as TNFRSF10B, FETUB, SCT, and ITGAV also contributed significantly to prediction outcomes. GAST and FETUB ranked among the highest contributors in several subsets, while CA14, ASAH2, and PZP emerged as prominent predictors in others (**Extended Data Fig. 6**). Including participants with cancer did not change the results, as CA14 remained the top-ranking protein (**Extended Data Fig. 6**).

### Prospective association of Top 15 proteins with pain phenotypes

We investigated the prospective associations between the levels of the top 15 proteins of the widespread chronic pain, top 15 proteins (CA14, ASAH2, TNFRSF10B, GDF15, SELENOP, CCL20, SCT, IL18R1, VSIG2, LEP, GAST, CELA2A, APOF, IL15, and LGALS3), and mechanism pain classification (nociceptive pain, neuropathic pain, nociplastic pain, and fibromyalgia), the onset, spread, and recovery of chronic pain.

Except for CELA2A, the other proteins exhibited significant associations with nociplastic pain or fibromyalgia. None of the 15 proteins showed significant associations with nociceptive pain or neuropathic pain, indicating that these 14 proteins can effectively distinguish nociplastic pain/fibromyalgia from nociceptive and neuropathic pain up to 9 years in advance (**Fig.6A**). At the 2022 follow-up, the onset and progression of chronic pain showed significant associations with the top 11 proteins, excluding SCT, GAST, CELA2A, and IL15. Among these, CA14, TNFRSF10B, GDF15, VSIG2, and LEP exhibited significant associations with all aspects of chronic pain onset and progression, including pain spreading and recovery (**Fig. 6A**). The detailed prospective associations between the top 15 proteins and pain mechanism classification (nociceptive pain, neuropathic pain, nociplastic pain, and fibromyalgia), as well as the onset and progression of chronic pain, including non-linear associations, are presented in the **Supplementary Fig.2-16**.

**Fig 6.**
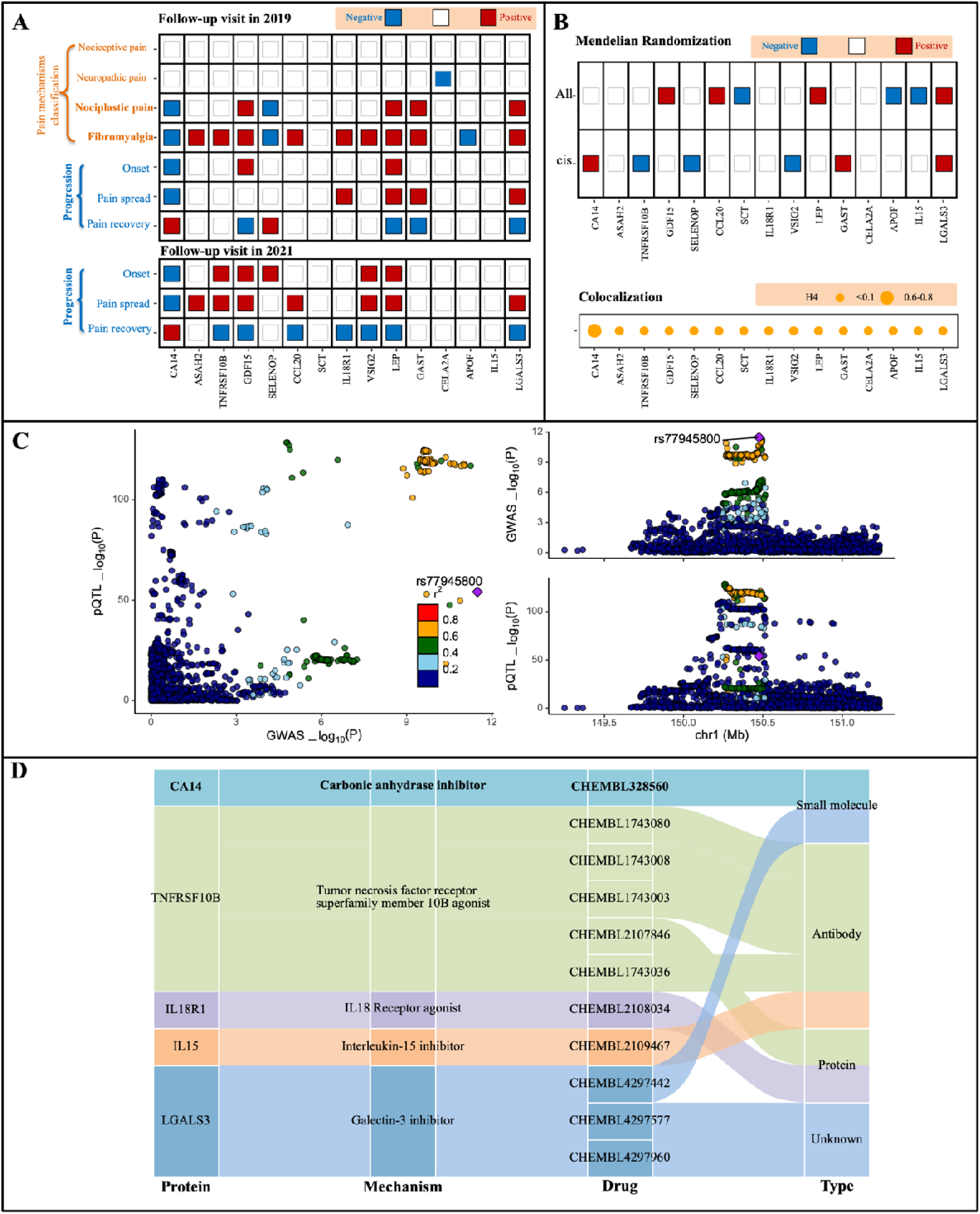
Prospective associations, mendelian randomisation results, and drug repurposing based on top 15 proteins and their potential causal relationships with chronic pain. Notes: Panel A: prospective associations between the top 15 proteins and chronic pain. The analysis was adjusted for several confounders, including sleeplessness, frustration, fatigue, mood disturbances, stressful life events, and body mass index (BMI). Prognostic associations for pain onset were estimated among participants who reported no pain at baseline. Panel B: mendelian randomisation and colocalisation results for the top 15 proteins. the colocalisation analysis utilised protein quantitative trait loci (pQTL) data from the UK biobank (UKB) to identify shared genetic variants between protein levels and pain outcomes. Mendelian randomisation was conducted to explore potential causal relationships between the top 15 proteins and multisite chronic pain. Panel C: colocalisation analysis for CA14 and multisite chronic pain. Panel D: identification of potential therapeutic targets based on the top 15 proteins.

### Mendelian Randomisation and Causal Inferences

Two-sample Mendelian randomisation (MR) analyses revealed causal effects of genetically predicted levels of 12 proteins (all but ASAH2, IL18R1, and CELA2A) and multisite chronic pain (**Fig. 6B** **and Supplementary Table 5**). Further colocalisation analyses reinforced the causal relevance of CA14 (H4 = 0.61) with multisite chronic pain (**Fig. 6C** **and Supplementary Table 6**). In the reverse MR, we found no significant association between genetic liability to multisite chronic pain and CA14. However, a significant association was observed with LEP (**Supplementary Table 6**).

The protein-protein interaction (PPI) network revealed that LEP, GDF15, and TNFRSF10B were central nodes, exhibiting multiple connections with other significant proteins such as LGALS3, IL15, and CCL20, suggesting their pivotal roles in chronic pain modulation. In contrast, proteins like CELA2A and APOF were more isolated, indicating potentially peripheral roles in the network (**Extended Data Fig. 7 and Supplementary Table 7**).

### Proteomics-based drug repurposing

Among the top 15 proteins, five (CA14, TNFRSF10B, IL18R1, IL15, and LGALS3) were found in the Open Targets database, highlighting their potential as therapeutic targets (Supplementary Table 8). A total of 11 drugs were identified targeting these proteins, including both small molecules and biologics such as antibodies and proteins, each categorised by drug type and mechanism of action. Notably, **CA14** emerged as a key therapeutic target, with sultiame (CHEMBL328660), a carbonic anhydrase inhibitor, as its main associated drug. SULTHIAME, which acts on the plasma membrane (**Extended Data Fig. 12**), is primarily used for treating obstructive sleep apnea and epilepsy. However, CA14 agonists may be a more suitable therapeutic approach, based on the non-linear association of the CA14. Beyond sulthiame, the remaining ten drugs present promising options for treating nociplastic pain

## Discussion

Our analysis revealed marked differences in the protein signatures of chronic versus acute pain. By leveraging these protein changes, we developed pain-specific proteomic scores that not only effectively discriminated individuals with chronic pain from those who were pain-free, but also predicted the intensity, phenotype, onset, spread, and recovery of chronic pain, particularly widespread chronic pain, a key feature of nociplastic pain. Notably, a sparse proteomic signature based on the top 10 proteins performed comparably to models based on conventional clinical variables alone. Among the top 15 proteins identified, 12 were significantly linked to these pain mechanisms, with 10 supported by Mendelian randomisation analysis. CA14, identified as the top-ranking protein and supported by both MR and colocalisation analyses, is a known target of sulthiame (CHEMBL328560), suggesting a potential therapeutic target for nociplastic pain conditions such as fibromyalgia.

### Distinct Protein Signatures in Chronic and Acute Pain

Chronic pain exhibits more extensive protein changes compared to acute pain. Traditionally, chronic pain is defined primarily by its duration and lacks specific biomarkers to guide clinical management. Our study addresses his challenge by identifying distinct proteomic signatures that differentiate between chronic and acute pain, suggesting that proteomic information could predict those who will transition from acute to acute pain. Previous research has highlighted the utility of proteomics in understanding pain sensitivity, especially in experimental models^18^. Notably, several proteins identified in our study provide novel insights into the biological pathways involved in pain mechanisms, in particular highlighting the involvement of metabolic and immune pathways in chronic pain development and persistence. For instance, leptin (LEP), known for its role in energy regulation, was strongly associated with chronic pain, indicating the potential involvement of metabolic dysregulation. Additionally, proteins such as ASGR1 and RARRES2, involved in immune responses, highlight the role of inflammation in chronic pain mechanisms. Inflammatory biomarkers may aid in predicting high-risk groups for developing nociplastic pain and guide the development of tailored treatments.

The distinct molecular profiles observed across specific pain types, such as head and face pain, abdominal pain, and musculoskeletal pain, further emphasise the heterogeneity of chronic pain. However, the identification of shared proteins across multiple pain types — such as APOF and ASGR1— suggests that there may be common underlying biological mechanisms driving pain across body regions. This finding could pave the way for the development of universal biomarkers that apply across multiple pain syndromes.

### Classification Accuracy of Proteomic Profiling

We demonstrate that proteomic profiling can perform as well as conventional clinical risk scores in predicting chronic pain and its progression. Moreover, combining proteomic scores with clinical variables further enhances predictive performance, suggesting that an integrated approach may be beneficial. This study is the first to comprehensively demonstrate the predictive power of plasma proteins across a broad spectrum of chronic pain types. We achieved this by directly comparing proteomic data with clinical variables and evaluating the added predictive value that proteomics offers beyond traditional clinical predictors. The proteomic score alone exhibited high predictive accuracy for chronic pain and its various subtypes. This predictive performance was further enhanced when proteomic data were combined with clinical variables. Our findings underscore the substantial potential of proteomic profiling as a promising technique to complement clinical evaluation, aiding in both the risk assessment for transition to chronic pain and the prognostication of chronic pain outcomes.

The proteomic score demonstrated a strong association with the onset, spread, and recovery of chronic pain. This profile effectively identifies individuals at higher risk for developing chronic pain, experiencing its spread to new sites, and their likelihood of recovery. Pain is influenced by a complex interplay of biological, psychological, and social factors ^13^. Proteins integrate genetic, environmental, age-related, behavioural, and medication influences, offering comprehensive insights into pain mechanisms. Previous research has suggested that pain spread is not random but shows strong dependencies between adjacent pain sites ^13^. Our study supports this by revealing significant associations between pain-specific proteomic scores and pain at various sites. Additionally, we observed robust correlations between proteomic scores and pain recovery, indicating shared biological foundations for both the spread and recovery of pain.

The proteomic score demonstrated a significant and specific association with nociplastic pain and fibromyalgia, distinguishing them from nociceptive and neuropathic pain. Nociplastic pain is characterised by abnormal sensory processing without clear peripheral damage. The association between proteomic score and nociplastic pain suggests that proteomic signatures capture the complex biological underpinnings of this condition, including abnormalities in sensory perception, a hallmark feature^11^.

### Causal Relevance and Therapeutic Potential of Key Proteins

Carbonic anhydrase 14 and leptin were identified as the primary proteins consistently associated with chronic pain, particularly nociplastic pain and fibromyalgia, and progression of chronic pain (onset, spreading, and recovery of chronic pain) in both longitudinal analyses. carbonic anhydrase 14 , identified here for the first time as associated with chronic pain, plays an important role in neuronal signal transmission via zinc ion binding and carbonate dehydratase activity. It is primarily enriched in the spinal cord and hasa crucial for neuronal function. Leptin is involved in immune regulation and inflammation, contributing to pain modulation, especially in nociplastic pain. Fibromyalgia affects 10–48% of patients with rheumatic diseases, compared to ∼2–6% in the general population, likely due to shared mechanisms of immune dysregulation and inflammation^19–21^. Both leptin and gastrin showed positive correlations with new-onset pain and pain spread, suggesting their role in the development and propagation of chronic pain. This relationship is further supported by their impact on pain recovery outcomes. Animal studies have demonstrated that leptin can influence pain thresholds ^22,23^. Together, these proteins suggest potential complementary roles in pain, with leptin influencing inflammation and carbonic anhydrase 14 affecting neuronal signalling.

The role of carbonic anhydrase 14 and leptin were both supported by mendelian randomisation, with carbonic anhydrase 14 further validated through colocalisation analysis. Our search on the Open Targets platform revealed that carbonic anhydrase 14 is the target of an approved small molecule drug, sulthiame, which is currently used to treat epilepsy and obstructive sleep apnoea. However, there are currently no well-documented drugs that activate CA14. Most research on carbonic anhydrase modulation has centered on inhibitors rather than activators, as seen with sulthiame in epilepsy treatment. If enhancing CA14 activity proves to be therapeutically relevant for pain management, future research will need to focus on developing specific agonist or allosteric modulators. In addition to carbonic anhydrase 14, drugs for four other proteins were identified, primarily used for cancer and autoimmune diseases. While these therapies carry potential risks related to immune modulation, they remain possibilities for treatment options. Carbonic anhydrase 14, the top-ranked protein with a strong causal association, suggests that targeting this protein could offer a promising therapeutic avenue for nociplastic pain.

### Limitations and strengths

While our study provides strong evidence for the role of proteomic profiling in characterisitng chronic pain, there are limitations which warrant discussion. First, while proteomic profiling demonstrated predictive value, the Olink panels used may have excluded key proteins relevant to chronic pain, potentially limiting the comprehensiveness of our analysis. Future studies should incorporate a wider array of proteomic markers to capture a more complete picture of the biological pathways involved in chronic pain. Second, the focus was primarily on a specific subset of pain types. Other pain characteristics not included may have different proteomic profiles and predictors. Furthermore, some participants in the control group, despite being classified as pain-free, may have had a history of chronic pain. Third, the study population was predominantly white British, which may limit the generalizability of the findings to other ethnic groups. Proteomic profiles and pain associations could vary across different genetic backgrounds. Fourth, while leptin and carbonic anhydrase 14 showed a causal relationship with pain, potential protein-protein interactions were not considered. The underlying biological pathways also remain unclear, warranting further research. Fifth, the findings have not been validated in independent cohorts. External validation in other large and diverse populations is necessary to confirm the robustness and applicability of the results. Finally, we acknowledge that the MR findings would be more robust if there were evidence indicating that the pQTLs and genetic variations associated with the outcome are shared. However, we also recognise that the lack of colocalisation evidence does not undermine the validity of the results, as colocalisation methods are known to have a high false-negative rate, typically around 60%^24^.

### Conclusion

In conclusion, this study underscores the significant potential of proteomic profiling to advance our understanding and management of chronic pain, particularly nociplastic pain and fibromyalgia. By identifying key proteins such as **carbonic anhydrase 14** and **leptin**, we offer new insights into biological mechanisms underlying pain. Therapies targeting TNFRSF10B, like tigatuzumab, represent promising candidates for drug repurposing in chronic pain treatment trials. Although no CA14 agonists are currently available, CA14 remains a highly promising target, encouraging future efforts to develop novel therapies that leverage its potential in pain modulation.

## Methods

### Study population

The UK Biobank is a prospective, population-based cohort comprising over 500,000 individuals aged 40-69, recruited from 22 assessment centers across the UK between 2006 and 2010. In 2019, additional online surveys were conducted to track specific pain-related outcomes over time. A random sample of participants (n=53,029) were selected for the measurement of proteomics data and thus included for the subsequent analyses. More details of study design, cohort construction, and study window is provided in **Extended Data Fig. 8** and **Extended Data Fig. 9**. Ethical approval was obtained from the North West Multi-centre Research Ethics Committee (MREC, https://www.ukbiobank.ac.uk/learn-more-about-uk-biobank/about-us/ethics), with all participants providing written informed consent. This research was approved under application number 98,358.

To reduce confounding effects from cancer diagnosis or treatment, participants with cancer-related pain were excluded, allowing the focus to remain on chronic non-cancer pain. Additionally, participants selected by the UK Biobank Pharma Proteomics Project (UKB-PPP) Consortium were excluded from the analyses.

### Clinical variables

Six variables capturing biological, psychological, and social factors were included as predictors for the construction of a clinical model. They were specified in prior with literature evidence suggesting their predictive value for chronic pain ^13^. The factors were collected by an standardised touchscreen questionnaire in UK Biobank and include sleep difficulties (difficulty falling asleep, rated as quite a lot or very much), neuroticism (perceived life effort as constant, rated as quite a lot or very much), powerlessness (feelings of lack of energy or powerlessness, rated as quite a lot or very much), mental health issues (history of diagnosis or treatment by a doctor), life stressors (events such as divorce, death of a partner, or significant unemployment history), and a BMI over 30kg/m^2^. All the variables listed above were transformed into binary variables.

### Proteomic profiling

The UK Biobank Pharma Proteomics Project consortium has generated extensive blood-based proteomic data. Proteomic profiling was conducted on EDTA-plasma samples from a cohort of 54,893 UKB participants at baseline recruitment, derived from a randomised subset representative of the broader UKB population. Utilising the Olink Explore 1536 and Expansion platforms, the profiling encompassed 2,923 unique proteins, assessed through 2,941 assays. The protein targeting assays are organised into four 384-plex panels:

Inflammation, Oncology, Cardiometabolic, and Neurology. Detailed methodologies concerning assay execution, sample selection, and handling are available in the online document (https://biobank.ndph.ox.ac.uk/showcase/label.cgi?id=1839). No notable impacts from batch and plate variations, nor anomalies in protein coefficients of variation (CVs), were detected. The inter- and intraplate CVs for all OLINK panels remained below 20% and 10%, respectively, with a median protein CV of 6.7%. There were strong correlations observed for identical proteins across different panels and between the OLINK assay and independent assays within the UK Biobank. The proteomics data were provided as Normalised Protein eXpression (NPX) values on a log_2_ transformation.

This analysis focuses on participants with less than 50% missing proteomic data. Additionally, three proteins missing in over 50% of participants were excluded. Post-quality control, missing NPX values were imputed using chained random forests, incorporating age and sex as predictors in the imputation process. The NPX data of proteins were standardised to have a mean of 0 and a variance of 1, following a standard normal distribution.

### Baseline pain phenotype

At the baseline recruitment between 2006 and 2010, 501,243 participants were asked about pain experiences that interfered with their usual activities in eight major body regions in the past one month.

Participants who reported pain at a specific site in the past month were subsequently asked whether the pain had persisted for more than three months. This follow-up question was used to define chronic pain (pain persisting for more than three months) and acute pain (having pain in last one month but persisting for three months or less). Among the eight painful locations specifically asked in the questionnaire, we grouped them into four categories based on the similarity of underlying aetiologies and conditions: (1) headache and facial pain, which may reflect conditions like migraines or tension headaches; (2) abdominal pain, potentially linked to visceral causes such as irritable bowel syndrome (IBS) or endometriosis; (3) musculoskeletal pain (neck or shoulder pain, hip pain, and knee pain), which may indicate localized mechanical or inflammatory issues; and (4) widespread pain (“pain all over the body”), often associated with conditions like fibromyalgia. For widespread pain, participants who reported experiencing it were not given the option to select other specific body regions. **Longitudinal Pain Phenotypes**

### Online UK Biobank in 2019 - Pain mechanism classification

Pain outcomes were classified based on participants’ chronic pain status and diagnostic criteria derived from validated scales. In 2019, participants were invited to complete an online pain questionnaire that incorporated the **2016 revised Fibromyalgia Survey Criteria (FSC)** ^25^and the **Douleur Neuropathique 4 (DN4)**^26^.

**Douleur Neuropathique 4 (DN4)**: The DN4 is a seven-item questionnaire designed to assess neuropathic pain. In the UK Biobank study, an abbreviated version was used, omitting the clinical examination assessment. Scores range from 0 to 7, with a score of ≥3 indicating the presence of neuropathic pain. Although primarily used to diagnose neuropathic pain, the DN4 may also reflect central sensitisation, a key feature of nociplastic pain^27^.

**Fibromyalgia Survey Criteria (FSC)**: The FSC is based on the 2016 ACR Fibromyalgia Survey Criteria^25^ and consists of two components: the **Widespread Pain Index (WPI)**, which measures pain across 19 body areas (score 0-19), and the **Symptom Severity Scale (SSS)**, which assesses somatic symptoms including fatigue, sleep disturbances, and cognitive difficulties (score 0-12). The total score (range 0-31) is calculated by combining the WPI and SSS scores.

The WPI was used to assess the severity of nociplastic pain, while the DN4 helped to identify both neuropathic pain and potential central sensitisation, providing a comprehensive evaluation of different pain types.

Based on these criteria, participants were classified into four distinct pain categories: **Nociceptive pain** was defined for participants with chronic pain but no neuropathic characteristics (DN4 score = 0) and an FSC score <3, indicating a low likelihood of nociplastic pain. Participants without chronic pain were classified as having no nociceptive pain.

**Neuropathic pain** was defined for participants with chronic pain and a DN4 score ≥ 3 suggesting the presence of neuropathic pain. Participants with an FSC score ≥ 3 were excluded to reduce the likelihood of including those with possible nociplastic or fibromyalgia-related pain.

**Nociplastic pain** was defined for participants exhibiting DN4 ≥3 or a total WPI and SSS score >12. Notably, universally accepted diagnostic criteria for nociplastic pain remain absent. Fibromyalgia, widely recognized as the prototypical manifestation of nociplastic pain, highlights the utility of the FSC score as an effective marker of symptom severity. In addition, the FSC score may reliably reflect the burden of nociplastic pain^28^. Accordingly, an FSC score ≥12 serves as a practical threshold for identifying individuals likely to experience nociplastic pain as the predominant driver of their chronic pain.

**Fibromyalgia** was identified either through self-reported diagnoses from baseline or the 2019 questionnaires or via general practitioner records. Additionally, participants with a WPI and SSS combined score >12 were classified as having fibromyalgia^28^.

**Online UK Biobank in 2019 - Pain sites:** Participants were inquired about experiencing pain or discomfort, either constantly or intermittently, for a duration exceeding three months. They were then requested to specify the affected sites, which included the head, face, neck, shoulders, chest, stomach, abdomen, back, hip, legs, knees, feet, arms, hands, and widespread body pain.

**New onset pain:** Participants who did not report any chronic pain at baseline but reported chronic pain at the follow-up visit were defined as having onset of pain.

**Spreading chronic pain:** The spreading of chronic pain was assessed by monitoring changes in the number of chronic pain sites. An increase in the number of pain sites at the follow-up visit compared to the baseline visit was defined as pain spread.

**Recovery of Chronic Pain:** Recovery of chronic pain was defined as a decrease in the number of pain sites at the follow-up visit compared to the baseline visit.

**Duration of Pain**: Participants reporting chronic pain were asked to specify the duration of their discomfort through the online questionnaire, selecting from the following categories: 3– 12 months, 1–5 years, or more than 5 years.

**Pain Rating in the Last 24 Hours:** Participants were also asked to rate their pain intensity at each reported chronic pain site over the preceding 24 hours using a numeric rating scale of 0 to 10, where 0 represented no pain and 10 indicated the worst possible pain.

**Worst Pain Rating**: Participants were prompted to rate the most severe pain they had experienced in the last 24 hours at the chronic pain site that bothered them the most, again on a numeric rating scale from 0 (no pain) to 10 (the worst imaginable pain). or widespread pain, a separate questionnaire (“All-over body pain in the last three months and rating of pain”) was used. The ratings from specific body sites and the overall widespread pain were combined for analysis.

**Pain Interference**: The impact of pain on daily functioning was evaluated across seven domains: general activity, mood, walking ability, normal work, relationships with others, sleep, and enjoyment of life. Each domain was rated on a scale of 0 to 10, where 0 indicated no interference and 10 signified complete interference. A total pain interference score was derived by summing the individual scores for these seven items.

**Symptom Severity**: Participants were asked to report the severity of three symptoms— fatigue, sleep quality, and cognitive symptoms—over the past week, as measured by the fibromyalgia symptom severity scale. Each symptom was rated on a scale from 0 (no problem) to 3 (severe, pervasive, continuous, and life-disrupting issues). The total symptom severity score was calculated by summing the scores for fatigue, sleep quality, and cognitive symptoms.

**Online UK Biobank in 2022- Pain sites:** In 2022, the UK Biobank conducted an online survey to assess pain at various anatomical sites among participants. Respondents were asked to identify current pain in specific areas, including the neck, back, chest, abdomen, legs, muscles, and joints. Additionally, participants provided information on the duration of the pain and the degree to which each site of pain impacted their daily lives. Pain persisting for more than twelve weeks was classified as chronic pain.

The onset of chronic pain, spreading chronic pain, recovery of chronic pain in the follow-up questionnaire 2022 was also defined followed the phenotype at follow-up questionnaire 2019.

## Statistical analyses

### Proteomic score

We employed a rigorous four-step machine learning framework (**Extended Data Fig.10**), which included: (1) feature selection through protein-wide association studies (PWAS), (2) development of a XGBOOST (Extreme Gradient Boosting) model for intricate proteomic scores (I-ProtS) based on all proteins with significant associations after Bonferroni correction, (3) development of a XGBOOST for sparse proteomic scores (S-ProtS) based on the top 10 proteins ranked by SHAP value, and (4) model validation. XGBoost is a high-performance machine learning algorithm designed for efficiency, flexibility, and portability, utilizing parallel tree boosting to develop the protein-profiling model^29^.

### Feature Selection

All participants were included in the feature selection phase. A protein-wide association study was conducted on 2,923 proteins using a binomial regression model with Bonferroni correction. Proteins with p-values less than Bonferroni-corrected p-value (0.05/2923), were selected for the development of the GBM (gradient boosting machines) to generate the pain-specific proteomics scores.

The proteins identified by the protein-wide association study, associated with specific chronic pain, represent the protein signature. To explore these protein signatures in distinguishing chronic from acute pain, acute pain — typically transient and driven by environmental factors — was used as a negative control to confirm the specificity of the protein signatures associated with chronic pain.

### Proteomic score development

We developed a series of XGBOOST models to assess the potential of plasma proteomics as a single-domain assay for predicting widespread chronic pain. For each pain type, a separate XGBOOST model was constructed using 10-fold cross-validation, a technique employed to minimise the risk of random error caused by data splitting. The predicted risk values from these models were designated as proteomic scores (intricate proteomic scores, I-ProtS). To identify the most important proteins contributing to the prediction of each pain type, we used SHAP (SHapley Additive exPlanations) values. SHAP values are derived from cooperative game theory and provide a unified measure to explain the contribution of each feature—in this case, each protein—to the model’s prediction. A positive SHAP value indicates that the protein increases the predicted risk for widespread chronic pain, while a negative SHAP value suggests the protein decreases the predicted risk. For each pain type, the top 15 proteins were ranked based on their SHAP values, indicating their relative importance in the model. SHAP plots were also generated to visually interpret the impact of each protein on the model’s predictions, allowing for a clear understanding of how specific proteins influence the risk predictions for widespread chronic pain.

Based on the top 10 proteins, we developed an additional XGBOOST model to generate a spare-proteomic-score (S-ProtS), focusing on a more compact set of features. This model aims to reduce complexity while maintaining predictive power, leveraging only the most important proteins identified from the previous analysis.

### Model for pain prediction

We fitted GBM models incorporating different sets of clinical predictors (**Extended Data Fig.11**). Specifically, 80% of participants were designated for training, while 20% were used for validation. We developed models based on six predictors identified from a previous study, utilizing 10-fold cross-validation. Additionally, we created models combining these six clinical predictors with proteomic score to evaluate their additive predictive value. Model performance was assessed using AUC, ranging from 0.5 (non-informative) to 1 (perfect discrimination). Calibration plots were employed to visualise the agreement between predicted risks and observed pain rates, while net benefit curves illustrated the additive predictive value of ProRS alongside different clinical predictor sets.

We reported key predictive metrics, including the area under the Receiver Operating Characteristic (ROC) curve (AUC). Calibration was evaluated using calibration plots and quantified through the calibration slope statistic, with a slope of 1 indicating optimal overall calibration. All models underwent development and evaluation through 10-fold cross-validation, with continuous variables standardised and categorical variables one-hot encoded. The results for all participants, including those with cancer, are provided as a sensitivity analysis in the supplementary materials to assess whether the presence of cancer affects the contribution of proteins.

### Prospective association of the proteomic score

To evaluate the prognostic association of the widespread chronic pain-specific proteomic score, Poisson regression was employed to estimate the relationship between the proteomic score (quantile 5 vs quantile 1) and nociceptive pain, neuropathic pain, nociplastic pain, fibromyalgia, onset chronic pain, pain spread, and pain recovery. For onset pain, the analysis focused on participants without any chronic pain at baseline who developed onset pain by the follow-up visit.

### Prospective association of the top proteins

The top 15 proteins were identified as those with significant contributions to widespread chronic pain. Poisson regression was employed to estimate the prognostic association between these top proteins and pain mechanism classification (nociceptive pain, neuropathic pain, nociplastic pain, and fibromyalgia), new onset chronic pain, pain spread, and pain recovery (quantile 5 vs quantile 1). The analysis was adjusted for variables including sleeplessness, feeling fed-up, tiredness, mood, stressful life events, and BMI.

To explore dose-response relationships between the top proteins and pain outcomes, restricted cubic spline (RCS) regression was used, with the same multivariable adjustments. Nonlinearity was tested using the likelihood ratio test, and the number of nodes was selected based on the lowest Akaike Information Criterion (AIC) value. The same analytical approach was applied to the other top 15 proteins.

### Causal relevance of the top proteins (Mendelian Randomisation Analysis)

To explore the causal relationship between specific proteins and pain, we conducted a two-sample Mendelian randomisation (MR) analysis. This study targeted the top 2 shared proteins frequently appearing among the top ten across pain types: leptin (LEP) and gastrin (GAST). Selection of genetic instruments: Instrumental variables were derived from two large-scale genome-wide association studies (GWAS): the UK Biobank Pharma Proteomics Project (UKB-PPP). 1. All SNPs MR Analysis: Single nucleotide polymorphisms (SNPs) significantly associated with the expression of specific proteins in blood were selected as instrumental variables (p < 5 × 10^D5^). To minimise the influence of strong linkage disequilibrium (LD) on the outcomes, SNPs with low weak LD (R² < 0.1) within a 10000 kb window, as estimated using the 1000 Genomes European panel, were included. Additionally, F-statistics > 10 were applied to ensure sufficient instrumental strength related to exposure traits. 2. Cis-SNPs MR Analysis: Cis-SNPs associated with the top shared plasma proteins at genome-wide significance (p < 5 × 10_5) and LD (R² < 0.1) were utilised as instrumental variables. Cis-SNPs were defined as those located within 5 Mb of the gene encoding the protein, with LD estimated based on the 1000 Genomes European panel. Furthermore, F-statistics > 10 were applied to ensure adequate instrumental strength associated with exposure traits. This analysis was considered the primary MR result due to its proximal relevance to the gene encoding the protein.

Reverse MR analysis: The reverse MR used SNPs associated with multisite chronic pain at p < 5 × 10^D8^. Only SNPs with R² < 0.001 within a 10,000 kb window were included, and all instruments had an F-statistic > 10.

### Protein source: UKB-PPP

Pain sources: To avoid potential bias arising from population diversity, we restricted the genetic background in our drug target MR analysis to participants of European descent. We extracted multisite chronic pain data from a large cohort of GWAS^30^, which included 73,082 cases and 105,474 controls from the latest UK Biobank datasets.

### Sensitivity analysis

The Steiger test was employed to assess potential reverse causality in the MR analysis. Additionally, the bidirectional Mendelian randomisation approach was used to explore the reverse causal relationship between multisite chronic pain and specific proteins. Associations between cis-pQTLs and outcomes were calculated as beta with corresponding confidence intervals (CIs) using the random-effects inverse-variance weighted (IVW) method. Furthermore, MR-PRESSO was utilised to detect outlier genetic variants. The MR-Egger intercept test was conducted to evaluate horizontal pleiotropy, and Cochrane’s Q value was calculated to assess heterogeneity in IVW estimators, with P < 0.05 indicating evidence of heterogeneity.

### Colocalisation analysis

We performed colocalisation analysis to determine whether the observed associations between the top shared proteins and multisite chronic pain were driven by linkage disequilibrium. For each locus, a Bayesian framework was applied to evaluate support for five mutually exclusive hypotheses: (1) no association with either trait; (2) association with trait 1 only; (3) association with trait 2 only; (4) both traits are associated but have distinct causal variants; and (5) both traits are associated and share the same causal variant. The analysis yielded posterior probabilities for each hypothesis (H0, H1, H2, H3, and H4). We assigned prior probabilities as follows: the SNP being associated with trait 1 only (p1) was set at 1 × 10__; the SNP being associated with trait 2 only (p2) at 1 × 10__; and the SNP being associated with both traits (p12) at 1 × 10__. Strong evidence of colocalisation was defined as a posterior probability for shared causal variants (PH4) ≥ 0.8, while medium evidence was indicated by 0.5 < PH4 < 0.8.

### Protein-protein interaction (PPI)

We performed protein-protein interaction (PPI) analysis for the top 15 proteins identified in our models using the **STRING database** (https://string-db.org/)^31^. STRING offers comprehensive interaction networks derived from experimental data, computational predictions, and known biological pathways. This analysis enabled us to examine potential interactions between the top proteins and to identify biologically relevant networks that may be involved in the pathogenesis of widespread chronic pain.

### Drug Repurposing

To explore the potential for drug repurposing, we searched the Open Targets platform ^32^ for the top 15 proteins identified in our analysis. Open Targets integrates data from multiple sources, including genetic associations, known drug interactions, and disease pathways, to evaluate the therapeutic potential of various proteins. This search enabled us to determine whether any of the top proteins were associated with existing drugs that could be repurposed for chronic pain treatment. By identifying proteins already targeted by approved or investigational drugs, we aimed to uncover opportunities for repurposing these drugs to address widespread chronic pain. This approach may offer faster and more cost-effective therapeutic options by leveraging drugs that have already undergone substantial testing for safety and efficacy in other conditions.

All modeling and statistical analyses were performed using R software (V4.3.0). Missing value imputation was conducted using the missRanger package. The XGBOOST and gradient boosting model (GBM) was implemented with the caret package, with calibration curves generated by the CalibrationCurve package and net benefit curves by the rmda package. The two-sample Mendelian randomisation analysis was conducted using the TwosampleMR package. Colocalisation analysis was carried out with the *coloc* package.

## Data Availability

All data produced in the present study are available upon reasonable request to the authors

